# Plasma and stool metabolomic biomarkers of non-alcoholic fatty liver disease in Argentina

**DOI:** 10.1101/2020.07.30.20165308

**Authors:** Flavia Noelia Mazzini, Frank Cook, John Gounarides, Sebastián Marciano, Leila Haddad, Ana Jesica Tamaroff, Paola Casciato, Adrián Narvaez, María Florencia Mascardi, Margarita Anders, Federico Orozco, Nicolás Quiróz, Marcelo Risk, Susana Gutt, Adrián Gadano, Celia Méndez García, Martin Marro, Alberto Penas-Steinhardt, Julieta Trinks

**Affiliations:** Instituto de Medicina Traslacional e Ingeniería Biomédica (IMTIB) – CONICET - Instituto Universitario del Hospital Italiano (IUHI) - Hospital Italiano de Buenos Aires (HIBA), Ciudad Autónoma de Buenos Aires, Argentina; Analytical Sciences & Imaging (AS&I) Department, Novartis Institutes for Biomedical Research (NIBR), Cambridge, MA, United States of America; Liver Unit of Hospital Italiano de Buenos Aires, Ciudad Autónoma de Buenos Aires, Argentina; Nutrition Department of Hospital Italiano de Buenos Aires, Ciudad Autónoma de Buenos Aires, Argentina; Liver Unit of Hospital Alemán, Ciudad Autónoma de Buenos Aires, Argentina; Chemical Biology & Therapeutics (CBT) Department, NIBR, Cambridge, MA, United States of America; Cardiovascular and Metabolic Disease Area, NIBR, Cambridge, MA, United States of America; Universidad Nacional de Luján, Departamento de Ciencias Básicas, Laboratorio de Genómica Computacional, Luján, Buenos Aires, Argentina

**Keywords:** Non-alcoholic fatty liver disease, Biomarkers, Metabolomic, Microbiome

## Abstract

**Background and Aims:** Non-invasive biomarkers are urgently needed to identify patients with non-alcoholic fatty liver disease (NAFLD) especially those at risk of disease progression. This is particularly true in high prevalence areas such as Latin America. The gut microbiome and intestinal permeability may play a role in the risk of developing NAFLD and NASH, but the mechanism by which microbiota composition disruption (or dysbiosis) may affect NAFLD progression is still unknown. Targeted metabolomics is a powerful technology for discovering new associations between gut microbiome-derived metabolites and disease. Thus, we aimed to identify potential metabolomic biomarkers related to the NAFLD stage in Argentina, and to assess their relationship with clinical and host genetic factors.

**Materials and methods:** Adult healthy volunteers (HV) and biopsy-proven simple steatosis (SS) or non-alcoholic steatohepatitis (NASH) patients were recruited. Demographic, clinical and food frequency consumption data, as well as plasma and stool samples were collected. SNP rs738409 (PNPLA3 gene) was determined in all volunteers. HPLC and flow injection analysis with MS/MS in tandem was applied for metabolomic studies using the MxP Quant 500 Kit (Biocrates Life Sciences AG, Austria). Significantly different metabolites among groups were identified with MetaboAnalyst v4.0. Bivariate and multivariate analyses were used to identify variables that were independently related to NAFLD stage. Forward stepwise logistic regression models were constructed to design the best feature combination that could distinguish between study groups. Receiver Operating Characteristic (ROC) curves were used to evaluate models’ accuracy.

**Results:** A total of 53 volunteers were recruited: 19 HV, 12 SS and 22 NASH. Diet was similar between groups. The concentration of 33 out of 424 detected metabolites (25 in plasma and 8 in stool) was significantly different among study groups. Levels of triglycerides (TG) were higher among NAFLD patients, whereas levels of phosphatidylcholines (PC) and lysoPC were depleted relative to HV. The PNPLA3 risk genotype for NAFLD and NASH (GG) was related to higher plasma levels of eicosenoic acid FA(20:1) (p<0.001). Plasma metabolites showed a higher accuracy for diagnosis of NAFLD and NASH when compared to stool metabolites. Body mass index (BMI) and plasma levels of PC aa C24:0, FA(20:1) and TG(16:1_34:1) showed high accuracy for diagnosis of NAFLD; whereas the best AUROC for discriminating NASH from SS was that of plasma levels of PC aa C24:0 and PC ae C40:1.

**Conclusion:** A panel of plasma and stool biomarkers could distinguish between NAFLD and NASH in a cohort of patients from Argentina. Plasma biomarkers may be diagnostic in these patients and could be used to assess disease progression. Further validation studies including a larger number of patients are needed.

## 1. INTRODUCTION

Non-alcoholic fatty liver disease (NAFLD) is a multisystem disease strongly associated with obesity, insulin resistance/type II diabetes mellitus, high blood pressure and dyslipidemia [1]. It represents a dynamic and progressive spectrum of liver disease encompassing simple fatty infiltration of the liver parenchyma (simple steatosis, SS), and fatty infiltration and inflammation (non-alcoholic steatohepatitis; NASH). Notably, NASH is not by itself a severe hepatic lesion, but it can progress toward cirrhosis, decompensated liver disease, and hepatocellular carcinoma [2].

It is now recognized that NAFLD is the most common cause of chronic liver disease around the world, representing a major nutritional concern because of the high worldwide prevalence of overweight and obesity. Moreover, NAFLD is rapidly becoming the most common indication for liver transplant. Currently, its global prevalence is estimated to be 25%, but higher rates are observed in South America (31%) [3,4]. The severity of NAFLD also may be greater among people of Native American ancestry, probably due to the higher prevalence in these populations of the high-risk G allele of the single nucleotide polymorphism (SNP) rs738409 in the PNPLA3 gene, which is robustly associated with susceptibility to the clinical progression of NAFLD [5,6].

In order to early prevent liver damage and to improve clinical outcomes, the present challenge is to distinguish between SS and NASH as the latter increases the likelihood of liver disease progression. In this regard, liver biopsy is the “gold standard” as it provides important diagnostic and prognostic information [7]; however, it remains a costly and invasive procedure with inherent risks. Thus, it cannot be used as a tool for periodical monitoring. In addition, the amount of retrieved tissue can influence the diagnosis; and, interobserver differences are frequently encountered [8]. Currently, several serological scores are used in the clinical practice to identify patients with advanced fibrosis, but they do not distinguish between steatosis and NASH. Besides, they exhibit some limitations as indeterminate results are observed in a high rate of patients (30%), many of these scores have been validated in hepatology units and not in the general population, and their specificity is lower in some age groups [9,10]. Therefore, there is a growing medical need to discover novel non-invasive biomarkers that can predict the initial stage and progression of liver disease over time in an accurate manner.

In the last decade, it was revealed that changes in gut microbiome composition (dysbiosis) and bacterial metabolism products derive in alteration of intestinal permeability and function, contributing to the pathogenesis of several diseases, including NAFLD. Altered gut microbiome and its derived metabolites can facilitate the development of hepatic steatosis in patients at risk of NAFLD. Furthermore, gut dysbiosis has been shown to be associated with changes in levels of serum metabolites related to NAFLD [11]. In fact, the liver receives a high percentage of its blood supply from the splanchnic district through portal circulation, which exposes it to gut-derived molecules [12]. Since then, several studies have provided abundant evidence of the considerable interplay between gut microbiota and microbiota-derived compounds and the development and progression of NAFLD and NASH [13,14].

Each human’s gut microbiome differs due to enterotypes, body mass index (BMI) level, exercise frequency, lifestyle and cultural and dietary habits [15]. In addition, the gut microbiome differs among ethnicities [16] and thus, it would be of interest to study the presence of patterns that may contribute to a higher NAFLD prevalence or different disease severity in Latin America, where data is still scarce.

In this regard, metabolomics analysis is a new technology to explore mechanisms of different diseases, including minimal changes in microbiome-derived metabolites, which provides ample information on new biomarker discovery, disease pathogenesis, diagnosis, and personalized treatment. This powerful tool allows for the study of small-molecular intermediates and products of metabolism [17]. The analysis of specific patterns of metabolic alterations associated with NAFLD can help in providing insight into its etiology and mechanisms, as well as to discover novel disease biomarkers.

Therefore, we aimed to identify microbiome-derived metabolites that could be useful as non-invasive biomarkers for NAFLD and NASH in patients from Argentina and to assess their relationship with clinical and host genetic factors, contributing to gut microbiome knowledge and its association with disease in Latin America.

## 2. MATERIALS AND METHODS

### 2.1. Recruited subjects and sample collection

In this case-control study, adult participants were recruited at Hospital Italiano de Buenos Aires and Hospital Alemán, in Buenos Aires city, Argentina.

Subjects with persistently elevated liver enzymes were assessed by hepatologists at both mentioned hospitals and NAFLD was confirmed by liver biopsy and using standard medical practice to rule out other liver conditions [18]. Exclusion criteria for NAFLD patients were: schistosomiasis, hepatitis B virus (HBV) or hepatitis C virus (HCV) infection or any liver disease other than NAFLD, anticipated need for liver transplantation within a year or complications of end-stage liver disease such as variceal bleeding or ascites; concurrent medical illnesses; and contraindications for liver biopsy. Recruited NAFLD patients were then divided into two groups: simple steatosis (SS) and non-alcoholic steatohepatitis (NASH).

Healthy volunteers (HV) were recruited at the Nutrition Department of Hospital Italiano de Buenos Aires after a healthy eating consultation. All individuals had normal clinical and hematological examinations. In addition, they had normal liver enzyme levels and did not have any record of liver disease by ultrasound, and therefore showed no indication for liver biopsy.

Exclusion criteria for all groups of subjects were: antibiotics, laxatives or probiotics consumption in the preceding 6 months, use of medications known to cause or exacerbate steatohepatitis, history of pelvic radiation exposure or chemotherapy, previous gastrointestinal surgery modifying the anatomy, history of chronic gastrointestinal disease, inflammatory bowel disease or any other gut infectious disease, dietary restrictions (such as vegetarians or vegans), consumption of more than 20g of alcohol/day for women and 30g of alcohol/day for men, illegal drug consumption, pregnancy or lactating state.

After signing an informed consent statement upon enrollment, each subject provided one stool sample and one fasting blood sample, underwent anthropometric measurements (height and weight) and completed a self-administered food frequency consumption questionnaire [19, 20]. The participants’ demographics and medical history were also reviewed and collected.

Each stool sample was frozen immediately after collection in the participants’ home freezer (−20°C). Within 24 hours, they brought the frozen sample to our lab in an insulated box with cooling elements. Samples were stored at −80°C until lyophilization. Each lyophilized stool sample was homogenized and aliquoted into 2ml cryotubes and shipped for metabolomics analysis to Novartis Institutes for Biomedical Research (NIBR) based in Cambridge, MA, USA.

Each fasting blood sample was collected in a tube with potassium-EDTA and processed immediately after blood extraction. Plasma was separated by centrifugation at 20-24°C for 10 minutes at 2500 xg, transferred into a new pre-cooled tube and vortexed. Aliquots of 500µl were placed into 2ml cryotubes and stored at −80°C until shipping to NIBR for metabolomics analysis. Peripheral blood mononuclear cells (PBMC) were isolated from the buffy coat and stored at −80°C.

This study was approved by the Ethics Committee on Research from the Hospital Italiano de Buenos Aires, and it was conducted according to the Declaration of Helsinki.

### 2.2. DNA extraction and determination of the SNP rs738409 (PNPLA3)

Genomic DNA was extracted from PBMC by using QIAamp® DNA Mini Kit (QIAGEN, GmbH, Hilden, Germany) following the manufacturer’s protocol. SNP rs738409 in PNPLA3 gene was amplified as previously described [6]. The PCR amplified fragments were bi-directionally sequenced using Big-Dye Termination chemistry system (Applied Biosystems, Life Technologies Corp., Foster City, CA, USA) and the sequencing chromatogram was examined by using BioEdit Sequence Alignment Editor version 7.1.3.0.

### 2.3. Metabolomic analysis

Metabolite analysis of plasma and stool samples was performed using the MxP Quant 500 kit (Biocrates Life Sciences AG, Innsbruck, Austria).

Plasma samples were analyzed directly on the kit without the need for extraction.

For stool samples, 100mg of lyophilized feces were mixed with 6 times the volume of either extraction buffer A [ethanol/phosphate buffer (85:15 v/v)] for lipidic compounds or buffer B [ethanol/phosphate buffer (20:80 v/v)] for hydrophilic metabolites. In each case, samples were then sonicated at 80V for 3 cycles of 30 seconds each, then homogenized in vortex at 4°C for 10 minutes and centrifuged at maximum speed for 2 minutes at 4°C. Supernatant was transferred to a new pre-cooled tube. All samples were dried-down overnight and re-suspended into 200ul of the either extraction buffer A or B. Ten-fold dilutions were made from aliquots of samples extracted with buffer A.

Finally, 10µl of each plasma and extracted stool sample were loaded and run in separate plates. MxP Quant 500 Column System was used in combination with SCIEX Triple Quad™ 6500+ mass spectrometer (SCIEX, Danaher Corporation, Washington, D.C., United States) in both positive and negative modes. Data acquisition was performed using specific mass transitions (MRM pairs) and the Biocrates® Met*IDQ*™ software (Biocrates Life Sciences AG, Innsbruck, Austria).

### 2.4. Data analysis

For descriptive statistics, medians and interquartile range (IQR), or absolute number and percentages were used. The statistical analysis of demographic, clinical, and human genetic data was performed by contingency tables using Chi-square test for categorical variables and Kruskal-Wallis and Mann-Whitney U test for continuous variables, using GraphPad Prism version 8.0.2 for Windows (www.graphpad.com). For food frequency data analysis, a Chi-square test with Monte Carlo estimation was performed using IBM SPSS (Statistical Package for Social Sciences) Statistics for Windows, version 22.0 (IBM Corp., Armonk, N.Y., USA). In all cases, significant differences were considered only for *p*<0.05.

Metabolites data was first filtered according to the plate quality control and the percentage of missing values. Metabolites with more than 30% of missing values were not considered for further analysis. For the remaining data, missing values were replaced with half of the minimum concentration value for each feature.

Analysis of metabolite concentration data was performed by means of MetaboAnalyst 4.0 (www.metaboanalyst.ca) [19]. A Kruskal-Wallis test was used where a false discovery rate (FDR) adjusted *p*-value (*q*-value) <0.05 was considered to be statistically significant within groups. Then, a Mann-Whitney U test was performed for one-to-one comparisons between the 3 groups and between HV and patients with NAFLD (SS + NASH), using IBM SPSS Statistics for Windows, version 22.0 (IBM Corp., Armonk, N.Y., USA).

In order to reduce the dimensionality of the data and visualize samples grouping, principal component analysis (PCA) were carried out with R version 3.6.2 [20].

A correlation matrix between metabolites and subjects’ characteristics with statistically significant differences among the three groups of study was built by means of Spearman’s correlation using MetaboAnalyst 4.0 (www.metaboanalyst.ca) [19].

In order to achieve a predictive signature capable of discriminating between HV and NAFLD (SS + NASH) and between SS and NASH, forward stepwise logistic regression models were constructed using R version 3.6.2 [20] after adjustment for the confounder effect of body mass index (BMI) on the stool and plasma data by means of ANCOVA test using IBM SPSS Statistics for Windows, version 22.0 (IBM Corp., Armonk, N.Y., USA). Plasma metabolites, stool metabolites and/or clinical parameters (BMI and PNPLA3 genotype) were used to design the best feature combination that could establish a predictive model for disease. Receiver Operating Characteristic (ROC) curves were used to evaluate the accuracy of these models.

The global performance of each biomarkers model was evaluated using the Area Under the Curve (AUC) and the determination of sensitivity and specificity at the optimal cut-off point defined by the minimum absolute difference between the sensitivity and specificity curves.

## 3. RESULTS

A total of 53 participants were included in this study: 19 HV and 34 NAFLD patients (12 SS and 22 NASH). Demographic, clinical and genetic data of all the study participants are shown in Table 1. BMI was significantly different between groups, being higher among NASH patients and lower among HV (*q* = 4.49e-06). Moreover, the genotypic frequency of rs738409 SNP in PNPLA3 gene showed significant differences between groups, being the risk GG genotype the most prevalent among NASH patients (*q* = 0.0198). Food frequency consumption data showed that none of the food groups evaluated in the self-administrated questionnaire showed significant differences in their consumption between the study groups (Table 2).

**Table 1.**
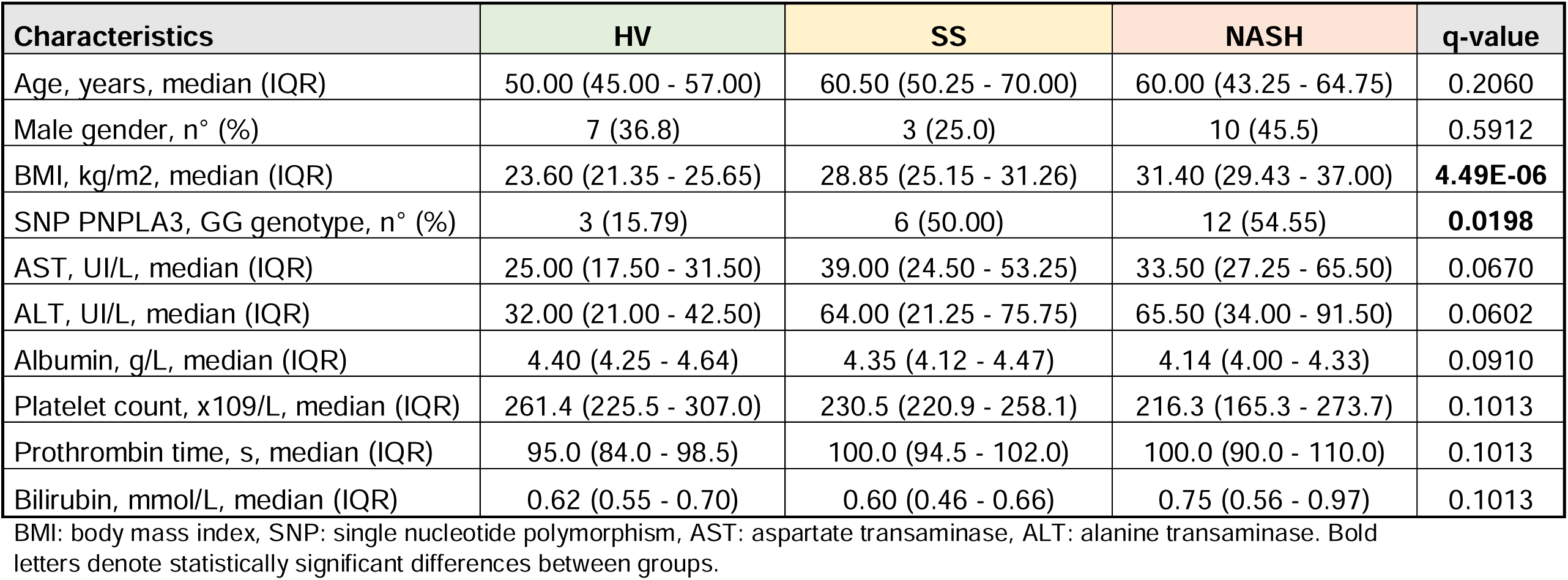
Demographic data, clinical characteristics and PNPLA3 genotypic frequency of the study subjects.

**Table 2.**
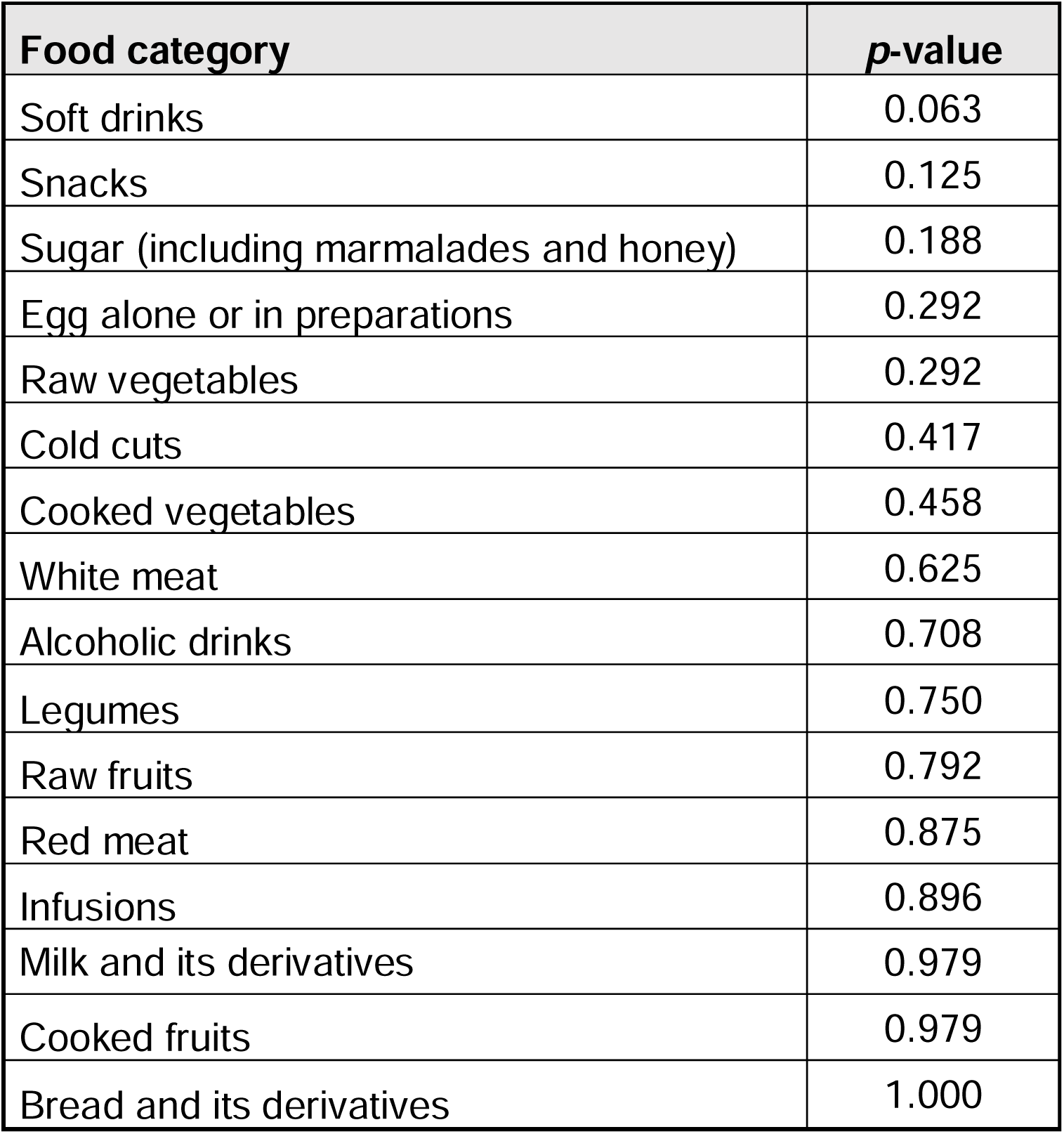
Food consumption frequency data analysis among groups.

Metabolomic analysis of plasma and stool samples detected a total of 458 metabolites at a quantifiable level, of which 424 passed data filtering: 229 metabolites were identified only in plasma samples, 24 only in stool samples and 171 in both sample types. Regarding the 3 groups of subjects, 1, 3 and 7 plasma metabolites were identified only among HV, SS and NASH patients, respectively (Supplementary Figure 1). In stool, after using extraction buffer A, 5, 4 and 10 metabolites were identified only among HV, SS and NASH patients, respectively (Supplementary Figure 1). However, extraction buffer B retrieved 3, 3 and 5 stool metabolites only detected among HV, SS and NASH patients, respectively (Supplementary Figure 1). The list of these metabolites is shown in Supplementary Table 1 and their category and annotation are detailed in Supplementary Table 2.

An initial PCA, based on the 424 metabolites that passed data filtering, identified sample N05 as an outlier (Figure 1). Visual inspection of the raw metabolomics data revealed that plasma triglycerides (TGs) levels in sample N05 were much higher than their concentrations in the other samples, which could probably be explained by the fact that this patient did not fast before the blood draw (Supplementary Figure 2). Thus, this sample was withdrawn from further analysis.

**Figure 1.**
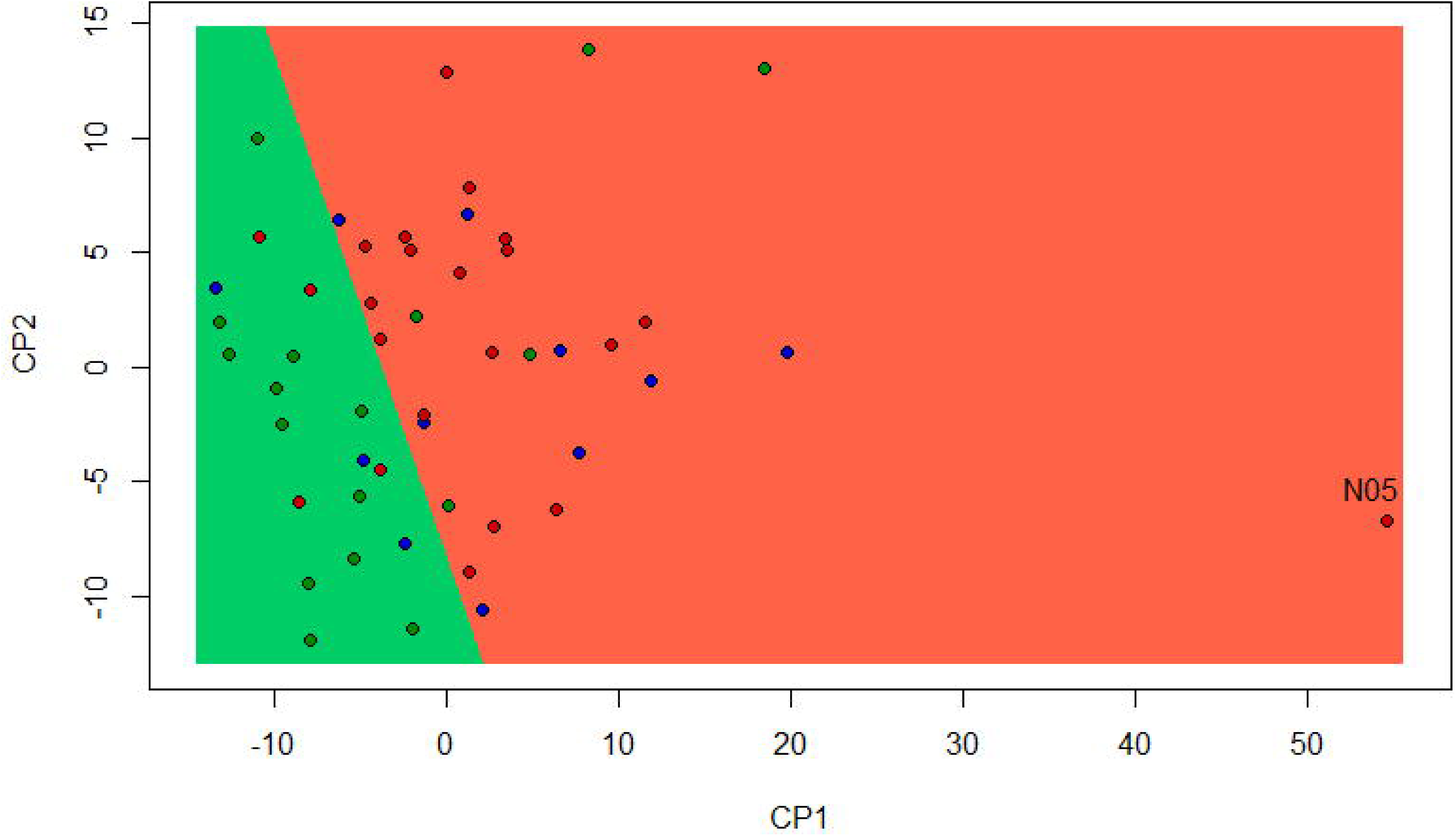
Principal component analysis (PCA) based on the 424 metabolites that passed data filtering in healthy volunteers (HV, green), patients with simple steatosis (SS, blue) and non-alcoholic steatohepatitits (NASH, red).

After analysis of the concentration of the detected metabolites, a total of 33 metabolites (25 in plasma and 8 in stool) significantly differed among groups (Table 3): all of them showed differences between HV and NASH patients; whereas 31 out of the 33 detected metabolites were significantly different between HV and NAFLD patients, and 15 metabolites showed different concentration levels between SS and NASH patients.

**Table 3.** Metabolites exhibiting statistically significant different concentrations among groups. Concentrations are expressed as median and interquartile range (IQR).

| Metabolite | HV | SS | NASH | q-value | One-to-one comparisons (*p*-value) |  |  |  |
| --- | --- | --- | --- | --- | --- | --- | --- | --- |
|  |  |  |  |  | HV - SS | HV - NASH | HV - NAFLD | SS - NASH |
| PLASMA |  |  |  |  |  |  |  |  |
| PC aa C24:0 | 0.112 (0.093 - 0.139) | 0.098 (0.087 - 0.114) | 0.064 (0.057 - 0.079) | **0.0004** | 0.2206 | **<0.0001** | **<0.001** | **<0.0001** |
| PC ae C40:1 | 1.22 (1.06 - 1.4) | 1.21 (1.045 - 1.29) | 0.929 (0.857 - 0.982) | **0.0086** | 0.9539 | **<0.0001** | **0.0060** | **0.0003** |
| PC ae C42:3 | 0.786 (0.675 - 1.01) | 0.73 (0.64 - 0.834) | 0.591 (0.546 - 0.642) | **0.0086** | 0.3290 | **<0.0001** | **<0.001** | **0.0120** |
| FA(20:3) | 1.02 (0.463 - 1.06) | 1.45 (0.7765 - 1.645) | 1.67 (1.34 - 2.45) | **0.0283** | 0.1153 | **<0.0001** | **<0.001** | 0.1042 |
| Glutamate | 24.3 (15.5 - 35.5) | 38.3 (26.05 - 65.95) | 45.5 (38.1 - 55.8) | **0.0283** | **0.0357** | **<0.0001** | **<0.001** | 0.4570 |
| Hippuric acid | 10.1 (6.29 - 16.6) | 20 (14.45 - 32.85) | 5.13 (2.33 - 6.47) | **0.0283** | **0.0221** | **0.0194** | 0.5220 | **0.0003** |
| lysoPC a C26:0 | 0.272 (0.213 - 0.364) | 0.22 (0.2035 - 0.2375) | 0.17 (0.126 - 0.211) | **0.0283** | 0.1246 | **0.0003** | **0.0020** | **0.0043** |
| lysoPC a C28:1 | 0.437 (0.333 - 0.493) | 0.312 (0.292 - 0.4415) | 0.264 (0.213 - 0.294) | **0.0283** | 0.1952 | **0.0002** | **0.0020** | **0.0086** |
| PC ae C36:2 | 21.2 (19.3 - 24.7) | 19.8 (17.75 - 22.75) | 15.7 (13.7 - 19.5) | **0.0283** | 0.3970 | **0.0002** | **0.0040** | **0.0086** |
| Tyrosine | 65 (48.5 - 73.8) | 71.5 (64.05 - 88.9) | 87.1 (76.5 - 99.5) | **0.0283** | 0.1872 | **<0.0001** | **0.0010** | **0.0449** |
| FA(20:1) | 1.49 (1.16 - 1.83) | 2.16 (1.75 - 2.63) | 2.41 (1.66 - 3.07) | **0.0321** | **0.0083** | **0.0002** | **<0.001** | 0.5313 |
| lysoPC a C28:0 | 0.283 (0.187 - 0.317) | 0.224 (0.178 - 0.2495) | 0.185 (0.106 - 0.189) | **0.0321** | 0.1027 | **0.0004** | **0.0020** | **0.0177** |
| TG(16:1_34:0) | 2.73 (2.06 - 6.11) | 8.54 (6.2 - 12.7) | 9.31 (6.51 - 11.3) | **0.0329** | **0.0056** | **0.0004** | **<0.001** | 0.5378 |
| TG(16:1_34:2) | 19.5 (10.5 - 29.6) | 38.1 (29.55 - 59.05) | 46.6 (31.8 - 60.3) | **0.0329** | **0.0123** | **0.0002** | **<0.001** | 0.6324 |
| TG(16:0_34:3) | 24.4 (13.9 - 31.9) | 41.6 (34.5 - 67.1) | 57.5 (35.1 - 67.2) | **0.0345** | **0.0109** | **0.0003** | **<0.001** | 0.6961 |
| TG(16:1_34:1) | 29 (17.3 - 53.7) | 64.3 (55.35 - 106.05) | 74.6 (60.2 - 91.3) | **0.0418** | **0.0109** | **0.0004** | **<0.001** | 0.7475 |
| Cystine | 55.7 (46.4 - 64.4) | 72.5 (54.9 - 79.4) | 86.5 (64.2 - 96.3) | **0.0444** | 0.0754 | **0.0004** | **0.0010** | 0.1114 |
| PC ae C40:6 | 5.03 (4.32 - 5.84) | 4.56 (4.225 - 5.405) | 3.69 (3.34 - 4.4) | **0.0444** | 0.6188 | **0.0023** | **0.0180** | **0.0048** |
| TG(16:0_32:0) | 10.2 (4.7 - 19.8) | 21.2 (16.55 - 35.6) | 26.5 (16.7 - 44.8) | **0.0444** | **0.0313** | **0.0005** | **0.0010** | 0.3725 |
| TG(16:0_34:2) | 71.1 (52.3 - 117) | 170 (121 - 231) | 186 (144 - 265) | **0.0444** | **0.0221** | **0.0005** | **0.0010** | 0.5249 |
| TG(16:1_34:3) | 4.14 (2.51 - 5.53) | 7.56 (5.13 - 12.5) | 8.42 (5.62 - 9.89) | **0.0444** | **0.0091** | **0.0006** | **<0.001** | 0.8993 |
| TG(20:4_32:0) | 2.5 (1.46 - 3.6) | 4.91 (2.56 - 7.555) | 5.76 (2.97 - 8.56) | **0.0444** | **0.0220** | **0.0005** | **0.0010** | 0.5637 |
| TG(16:1_32:1) | 6.41 (4.55 - 11.5) | 16.9 (11.25 - 23.75) | 15.5 (10.5 - 24.3) | **0.0453** | **0.0199** | **0.0005** | **0.0010** | 0.8452 |
| TG(16:0_34:0) | 8.46 (7.24 - 18.7) | 20.7 (13.55 - 34.8) | 24.8 (19.1 - 39.5) | **0.0474** | **0.0378** | **0.0006** | **0.0010** | 0.4050 |
| TG(16:1_32:0) | 4.64 (2.87 - 8.34) | 11.1 (6.945 - 18.05) | 12.3 (6.73 - 31.2) | **0.0474** | **0.0216** | **0.0010** | **0.0010** | 0.3075 |

| STOOL (ext. buf. A) |  |  |  |  |  |  |  |  |
| --- | --- | --- | --- | --- | --- | --- | --- | --- |
| Cer(d18:1/18:0) | 0.124 (0.021 - 0.176) | 0.174 (0.14 - 0.4175) | 0.309 (0.223 - 0.5445) | **0.0299** | **0.0465** | **0.0001** | **0.0010** | 0.1968 |
| Glycine | 2.35 (2.35 - 6.46) | 6.54 (2.35 - 7.99) | 13.85 (6.5425 - 18.45) | **0.0299** | 0.2584 | **0.0001** | **0.0020** | **0.0315** |
| Xanthine | 8.13 (3.43 - 13.9) | 15.7 (5.48 - 24.45) | 36.25 (15.475 - 57.8) | **0.0299** | 0.1469 | **0.0002** | **0.0010** | **0.0387** |
| Spermidine | 2.38 (1.49 - 3.42) | 1.47 (0.7195 - 6.53) | 7.455 (4.0675 - 9.8925) | **0.0318** | 0.8621 | **0.0005** | **0.0140** | **0.0202** |
| DCA | 36.1 (18.1 - 50) | 53.6 (41.75 - 73.3) | 68.35 (48.65 - 84.725) | **0.0318** | **0.0296** | **0.0007** | **0.0010** | 0.2976 |
| GDCA | 0.156 (0.093 - 0.285) | 0.347 (0.2775 - 0.4195) | 0.4895 (0.34075 - 1.1125) | **0.0318** | 0.1107 | **0.0004** | **0.0020** | 0.1447 |
| Cysteine | 0.802 (0.652 - 1.21) | 0.604 (0.5005 - 0.7345) | 1.375 (1 - 1.8675) | **0.0318** | 0.2392 | **0.0067** | 0.1810 | **0.0025** |
| CDCA | 34.7 (18.9 - 50.2) | 54.4 (41.55 - 75.9) | 68.1 (48.975 - 88.275) | **0.0365** | **0.0287** | **0.0010** | **0.0010** | 0.3587 |
| STOOL (ext. buf. B) |  |  |  |  |  |  |  |  |
| Xanthine | 181 (89.85 - 278.5) | 160 (118.85 - 563.5) | 597 (378 - 748) | **0.0184** | 0.2871 | **<0.0001** | **<0.001** | **0.0325** |

HV: healthy volunteers, SS: simple steatosis patients, NASH: non-alcoholic steatohepatitis patients, NAFLD: non-alcoholic fatty liver disease.
Bold letters denote statistically significant differences between groups ( $p$ -value < 0.05).

The majority of the metabolites observed in plasma were lipidic compounds (84%) including TGs (44%), lysophosphatidylcholines (lysoPCs; 12%), phosphatidylcholines (PCs; 20%) and fatty acids (FAs; 8%), whereas the rest of the plasma metabolites were amino acids (8%), carboxylic acids (4%) and amino acid related compounds (4%). In stool samples, the detected metabolites were mostly bile acids (37.5%), but amino acids (25%), ceramides (12.5%), biogenic amines (12.5%) and nucleobase related compounds (12.5%) were also identified (Table 3). The list with the category and annotation of these metabolites are detailed in Supplementary Table 2.

A second PCA based on these 33 significantly different metabolites, showed a clear separation of samples in 3 groups, confirming that these metabolites could distinguish between the groups of recruited subjects (Figure 2).

**Figure 2.**
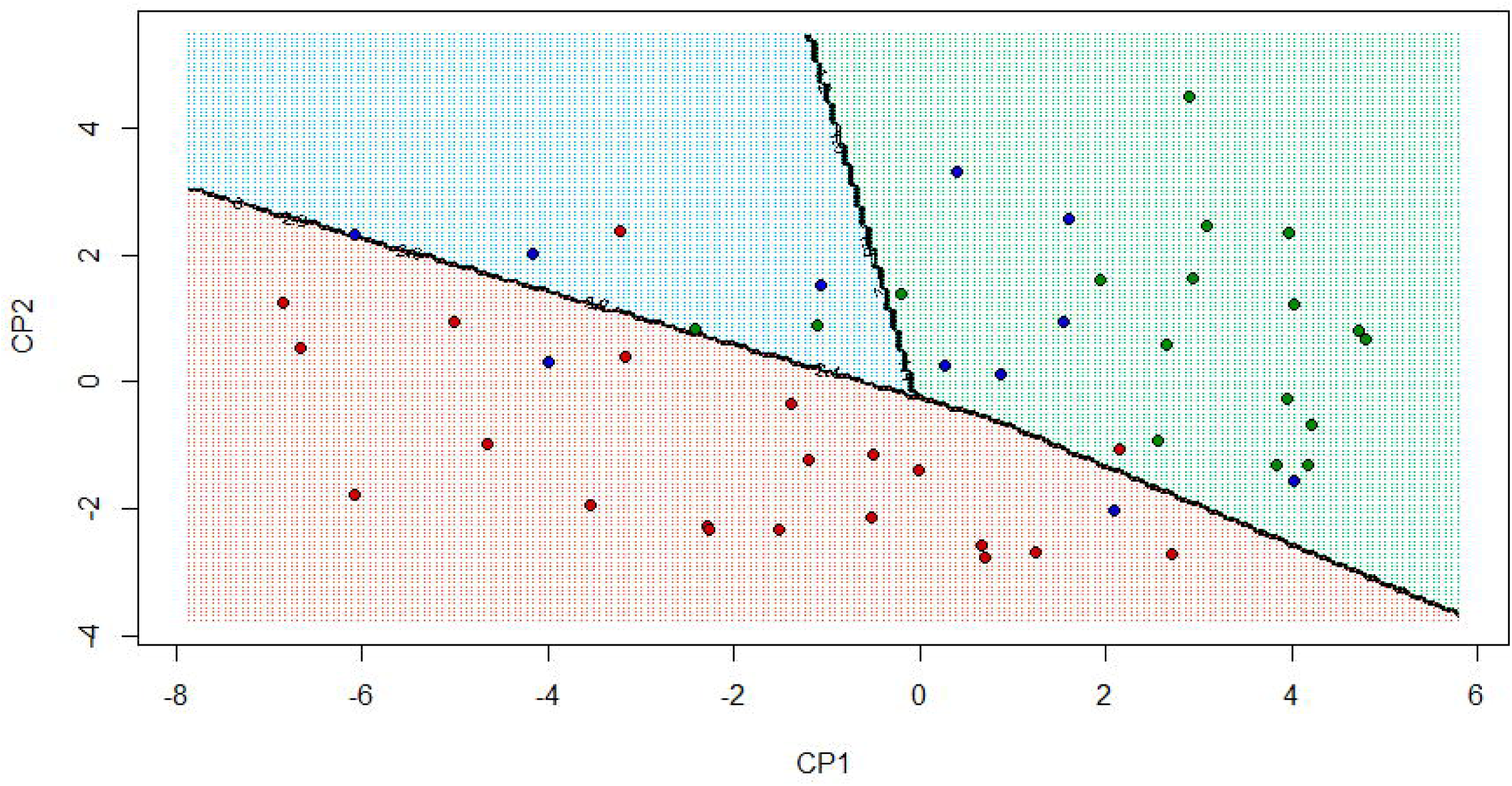
Principal component analysis (PCA) based on the 33 metabolites exhibiting statistically significant different concentrations among groups. Healthy volunteers, simple steatosis and non-alcoholic steatohepatitis patients are shown in green, blue and red, respectively.

In regard to these 33 metabolites, concentrations of plasma TGs, FAs and amino acids as well as the 8 identified stool metabolites were higher among NASH patients when compared to the other groups. In contrast, concentrations of plasma lysoPCs, PCs and the carboxylic acid hippuric acid were higher among HV (Table 3). In fact, analysis of Spearman’s rank correlation coefficients (ρ) confirmed these results: plasma lysoPCs and PCs were strongly negatively correlated with stool metabolites, whereas plasma TGs were positively correlated with them (Figure 3 and Supplementary Table 3).

**Figure 3.**
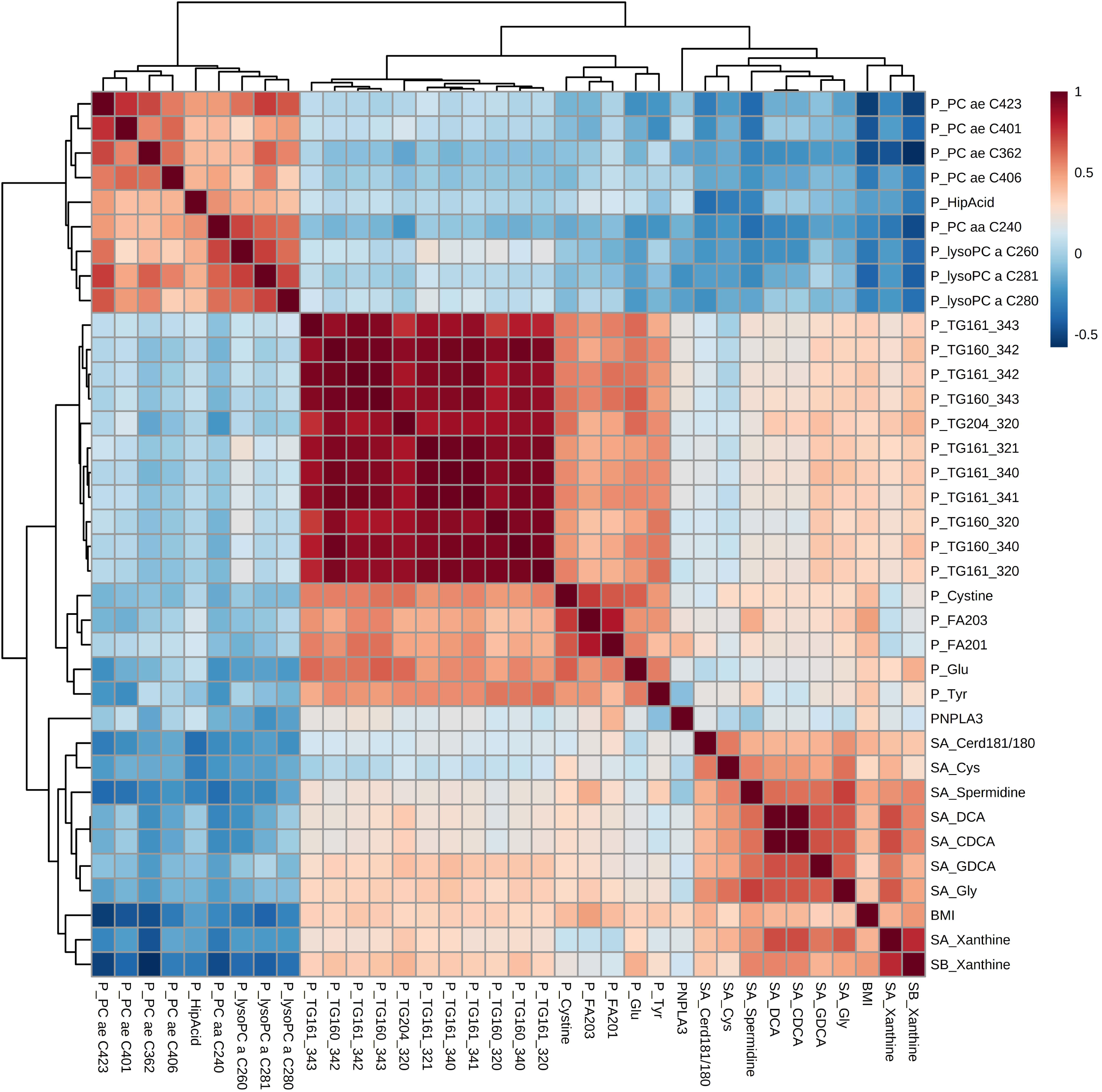
Heatmap of the Spearman correlation coefficients between plasma and stool metabolites, BMI and PNPLA3 genotype. Prefixes “P_”, “SA_” and “SB_” indicate those metabolites measured in plasma, or in stool samples extracted with buffer A and B, respectively.

The Spearman rank correlation analysis also showed a significant relationship between the PNPLA3 gene and the plasma levels of an eicosenoic acid, FA(20:1) (ρ=0.4240; *q*=0.035; Figure 3 and Supplementary Table 3). Further analysis revealed that the risk GG genotype is not only the most frequent among NASH patients, but it is also associated with higher plasma concentrations of FA(20:1); whereas lower concentrations of this metabolite were found among patients exhibiting the beneficial CC genotype for the aforementioned SNP (*p*=0.0007; Figure 4).

**Figure 4.**
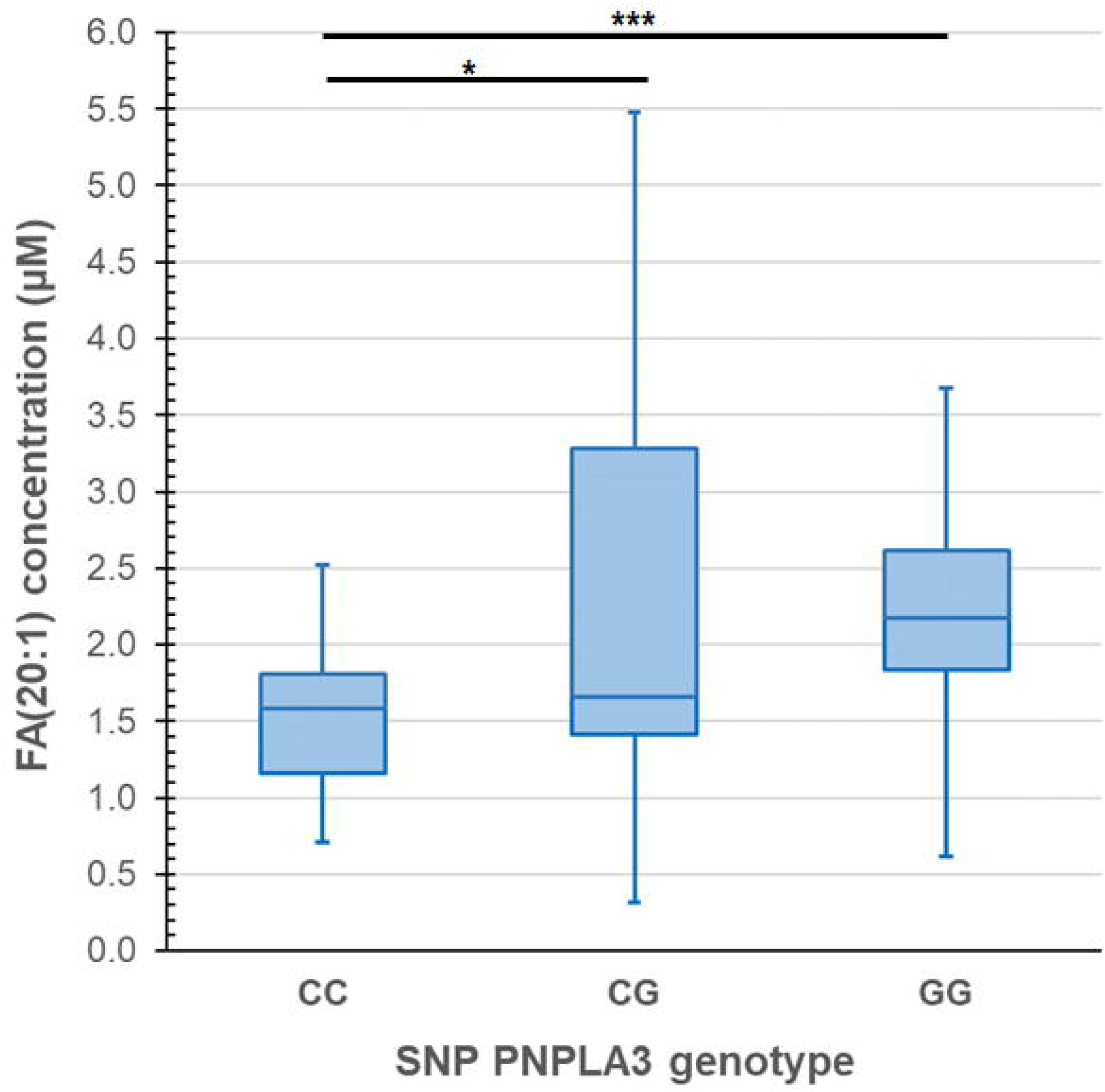
Association between the SNP rs738409 genotype in PNPLA3 gene and FA(20:1) concentration in plasma. \*\*\**p*=0.0007 and \**p*=0.0811.

Because NAFLD is closely linked to obesity, the relationship between the concentration of plasma and stool metabolites and NAFLD progression was also analyzed considering BMI as a covariate. The results showed that 24 metabolites (22 in plasma and 2 in stool) remained statistically significant (Table 4). The 19 metabolites with significant differences between HV and NAFLD patients were detected in plasma except for stool xanthine, being PC aa C24:0 the most significant one (Table 4). In the case of SS and NASH patients, 10 metabolites (8 in plasma and 2 in stool) exhibited significant differences between these groups of patients with dissimilar NAFLD stages, being PC aa C24:0 the most significant metabolite, as well (Table 4).

**Table 4.** Body mass index (BMI)-adjustment of significant metabolites’ *p*-values by ANCOVA test.

| Metabolite | *p*-value |  | BMI-adjusted *p*-value |  |
| --- | --- | --- | --- | --- |
|  | HV - NAFLD | SS - NASH | HV - NAFLD | SS - NASH |
| PLASMA |  |  |  |  |
| PC aa C24:0 | **<0.001** | **<0.0001** | **0.001** | **<0.001** |
| PC ae C40:1 | **0.0060** | **0.0003** | 0.600 | **0.003** |
| PC ae C42:3 | **<0.001** | **0.0120** | 0.061 | 0.056 |
| FA(20:3) | **<0.001** | 0.1042 | 0.102 | 0.197 |
| Glutamate | **<0.001** | 0.4570 | **0.002** | 0.556 |
| Hippuric acid | 0.5220 | **0.0003** | 0.275 | **0.004** |
| lysoPC a C26:0 | **0.0020** | **0.0043** | **0.021** | **0.037** |
| lysoPC a C28:1 | **0.0020** | **0.0086** | 0.188 | 0.097 |
| PC ae C36:2 | **0.0040** | **0.0086** | 0.287 | **0.018** |
| Tyrosine | **0.0010** | **0.0449** | **0.005** | **0.019** |
| FA(20:1) | **<0.001** | 0.5313 | **0.005** | 0.393 |
| lysoPC a C28:0 | **0.0020** | **0.0177** | **0.003** | **0.004** |
| TG(16:1_34:0) | **<0.001** | 0.5378 | **0.008** | 0.473 |
| TG(16:1_34:2) | **<0.001** | 0.6324 | **0.008** | 0.967 |
| TG(16:0_34:3) | **<0.001** | 0.6961 | **0.011** | 0.993 |
| TG(16:1_34:1) | **<0.001** | 0.7475 | **0.009** | 0.905 |
| Cystine | **0.0010** | 0.1114 | **0.043** | 0.267 |
| PC ae C40:6 | **0.0180** | **0.0048** | 0.498 | **0.022** |
| TG(16:0_32:0) | **0.0010** | 0.3725 | **0.009** | 0.262 |
| TG(16:0_34:2) | **0.0010** | 0.5249 | **0.016** | 0.679 |
| TG(16:1_34:3) | **<0.001** | 0.8993 | **0.010** | 0.452 |
| TG(20:4_32:0) | **0.0010** | 0.5637 | **0.019** | 0.520 |
| TG(16:1_32:1) | **0.0010** | 0.8452 | **0.013** | 0.532 |
| TG(16:0_34:0) | **0.0010** | 0.4050 | **0.011** | 0.402 |
| TG(16:1_32:0) | **0.0010** | 0.3075 | **0.034** | 0.141 |
| STOOL (ext. buf. A) |  |  |  |  |
| Cer(d18:1/18:0) | **0.0010** | 0.1968 | 0.140 | 0.766 |
| Glycine | **0.0020** | **0.0315** | 0.178 | 0.160 |
| Xanthine | **0.0010** | **0.0387** | 0.097 | 0.052 |
| Spermidine | **0.0140** | **0.0202** | 0.562 | 0.364 |
| DCA | **0.0010** | 0.2976 | 0.093 | 0.319 |
| GDCA | **0.0020** | 0.1447 | 0.363 | 0.326 |
| Cysteine | 0.1810 | **0.0025** | 0.924 | **0.011** |
| CDCA | **0.0010** | 0.3587 | 0.093 | 0.339 |
| STOOL (ext. buf. B) |  |  |  |  |
| Xanthine | **<0.001** | **0.0325** | **0.023** | **0.035** |

HV: healthy volunteers, SS: patients with simple steatosis, NASH: patients with non-alcoholic steatohepatitis, NAFLD: patients with non-alcoholic fatty liver disease.
Bold letters denote statistically significant differences between groups (*p*-value < 0.05)

To validate the importance of these metabolites, and to further gauge their ability to distinguish among patients with NASH and NAFLD and healthy controls, several diagnostic models were established and their potential predictive utility for the process of NAFLD was assessed by ROC curve analysis (Table 5 and Supplementary Figure 3). The predictors for NAFLD and NASH identified by logistic regression modelling are summarized in Figure 5.

**Table 5.**
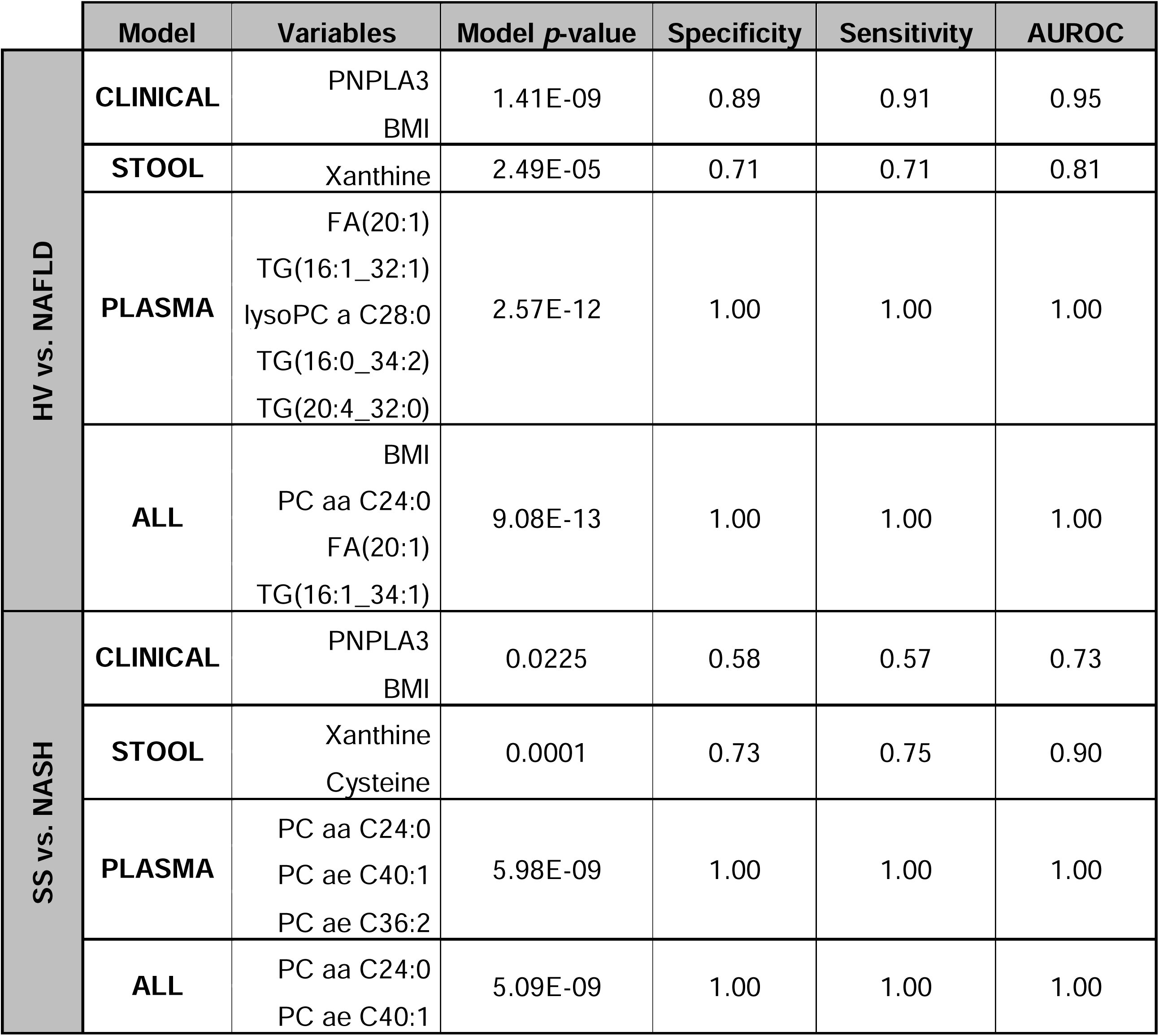
Diagnostic performance of different models for NAFLD and NASH.

|  | Model | Variables | Model *p*-value | Specificity | Sensitivity | AUROC |
| --- | --- | --- | --- | --- | --- | --- |
| HV vs. NAFLD | CLINICAL | PNPLA3<br>BMI | 1.41E-09 | 0.89 | 0.91 | 0.95 |
|  | STOOL | Xanthine | 2.49E-05 | 0.71 | 0.71 | 0.81 |
|  | PLASMA | FA(20:1)<br>TG(16:1_32:1)<br>lysoPC a C28:0<br>TG(16:0_34:2)<br>TG(20:4_32:0) | 2.57E-12 | 1.00 | 1.00 | 1.00 |
|  | ALL | BMI<br>PC aa C24:0<br>FA(20:1)<br>TG(16:1_34:1) | 9.08E-13 | 1.00 | 1.00 | 1.00 |
| SS vs. NASH | CLINICAL | PNPLA3<br>BMI | 0.0225 | 0.58 | 0.57 | 0.73 |
|  | STOOL | Xanthine<br>Cysteine | 0.0001 | 0.73 | 0.75 | 0.90 |
|  | PLASMA | PC aa C24:0<br>PC ae C40:1<br>PC ae C36:2 | 5.98E-09 | 1.00 | 1.00 | 1.00 |
|  | ALL | PC aa C24:0<br>PC ae C40:1 | 5.09E-09 | 1.00 | 1.00 | 1.00 |

**Figure 5.**
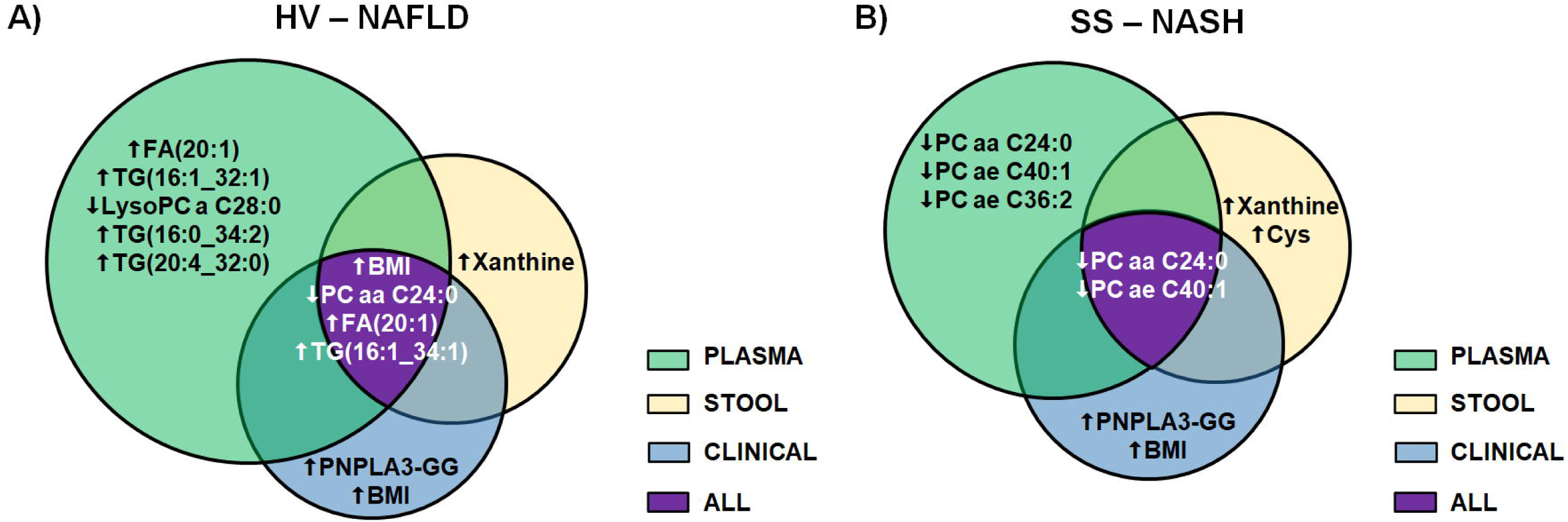
Predictors for NAFLD (A) and NASH (B) identified by logistic regression modelling where each Venn diagram represents the input data used for analysis. Significant variables identified by each model are displayed inside the corresponding diagram. Up-arrows designate variables that may possibly act as risk factors for NAFLD and/or NASH and down-arrows show variables that may be associated to a protective effect against the disease.

It is assumed that the AUROC of a model must be equal to or greater than 0.800 to be considered a less invasive test as good as a liver biopsy to evaluate liver damage [21]. Under this assumption, all models from the resulting analysis showed strong AUROC values, although the model that includes stool metabolites exhibited the lowest AUROC for NAFLD, and the clinical model, that includes PNPLA3 genotype and BMI, showed the weakest predictive power for NASH. For this latter model, the accuracy for discriminating NASH was lower than that for discriminating NAFLD; whereas the inverse was true for the stool model (Table 5 and Supplementary Figure 3).

When compared to stool metabolites, plasma metabolites showed higher accuracy for the diagnosis of NAFLD and NASH with AUROC values of 1.00 (Table 5 and Supplementary Figure 3). Modelling with all the significant features from our study (ALL model) revealed that the best AUROC was that of BMI and plasma levels of PC aa C24:0, FA(20:1) and TG(16:1_34:1) for diagnosis of NAFLD; whereas the best AUROC for discriminating NASH from SS was that of plasma levels of PC aa C24:0 and PC ae C40:1 (Table 5 and Supplementary Figure 3).

## 4. DISCUSSION

The microbiome and its derivatives represent a niche to be explored for NAFLD biomarkers discovery as its role in the pathogenesis of disease has already been described [11]. In this regard, several targeted metabolomic approaches had been carried out in the last years, most of them focusing only on lipidomics or analyzing just a few metabolites [22]. However, as microbiome is highly modified by several factors including ethnicity, diet and lifestyle [15,16], the obtained results are not necessarily applicable worldwide. Each population may have a personalized signature of NAFLD biomarkers, which makes it necessary to develop local studies, especially in South America where prevalence of disease is the highest [4]. In this study, we aimed to identify potential non-invasive, microbiome-derived biomarkers for NAFLD and NASH patients in Argentina. Therefore, we studied a wide range of metabolites in plasma and stool samples and established relationships with clinical and host genetic factors.

The recruited volunteers were residents of the same urban geographical area and were divided into 3 age and gender-matched groups. Considering that diet has been described as a modulator in NAFLD progression and one of the multiple factors that could modify the gut microbiota, and consequently its derived metabolites [23,24], we analyzed the food frequency consumption data from the recruited volunteers. Although it is recommended that NAFLD patients follow a healthy diet [25], our results showed no differences regarding food intake. For this reason, diet was not a confounding factor to consider within our cohort and all the reported changes in metabolites concentration among groups are independent of dietary factors.

The GG genotype of rs738409 SNP in PNPLA3 gene was the most frequent among NASH patients in our cohort. This risk genotype, which has been associated with severity and progression of NAFLD [26], showed a frequency of 55% among the NASH group. This prevalence rate is similar to those reported by other studies in admixed populations and could possibly explain the higher prevalence of NAFLD in Latin America populations [6,26–28].

In our cohort, BMI was significantly higher among NASH patients when compared to SS patients and HV. Being obesity a risk factor for NAFLD, it is expected that patients with disease have higher BMIs [29], although NAFLD can also manifest in lean patients [30]. Therefore, BMI could be considered as a confounding factor in our study.

The remaining analyzed clinical characteristics revealed no differences between the study groups. Although these variables are usually altered in NAFLD, normal levels of liver enzymes and albumin have been demonstrated in subjects within the entire spectrum of NAFLD, and therefore these clinical factors have not been very useful in predicting NAFLD [31,32].

There are multiple factors that may contribute to NAFLD development: gut microbiota dysbiosis, fat accumulation -which occurs simultaneously with toxic metabolites generation-, lipotoxicity and liver injury [12,33,34]. In our study, metabolomic analysis revealed a few metabolites uniquely identified in each study group, which could be useful as potential diagnostic markers. Acylcarnitines, which were associated with higher abundance of facultative anaerobes linked to inflammatory bowel disease [35,36] were distinctively detected in plasma and stool samples of NAFLD patients. Also, histamine was uniquely detected in NASH patients suggesting the presence of inflammatory response as it has been previously reported in other diseases [37].

Moreover, several metabolites associated with kidney function, such as creatinine were uniquely detected in NASH, implying a relationship between NAFLD and kidney disease [38]. However, further validation studies with a larger number of patients are needed to confirm these findings.

Quantitative analysis of metabolomic data initially revealed 33 metabolites that were able to discriminate between groups; however, after BMI-adjustment, 24 metabolites showed statistically significant differences. In agreement with previous studies [39,40], plasma levels of TGs, FAs and amino acids were increased in SS and NASH patients, whereas plasma concentrations of PCs and LysoPCs were decreased relative to HV.

Pathogenesis of NAFLD may be favored by higher levels of free FAs, causing inflammation, and leading to fat accumulation [41]. Moreover, long-chain FAs had been related to increased levels of pathogenic bacteria in the gut, contributing to inflammation and increased permeability [42].

Increased levels of circulating TGs in NAFLD patients has been also described in patients with obesity, diabetes or insulin resistance which are risk factors for this disease [29]. In NAFLD, a direct relationship between gut microbiota and TGs levels was described as probiotics treatment decreases TGs levels and intrahepatic fat [34,43]. Moreover, liver TGs accumulation could also be a consequence of lower concentrations of PCs in plasma [44,45]. In fact, choline is the principal precursor for PCs and LysoPCs synthesis, and its deficiency was associated with increased microbiota metabolism that could decrease host concentration levels of PC and LysoPC in NAFLD and NASH [13,46].

Hippuric acid is another plasma metabolite that showed significant differences between groups, being higher in subjects with SS than in HV and NASH patients. This carboxylic acid is a toxin derived of intestinal bacterial fermentation that may contribute to liver toxicity and lipid accumulation, and could be an early biomarker for NAFLD contributing to initial stages of disease progression [47,48].

Amino-acids metabolism has also been described to be modified in NAFLD. Consistently with our results, higher glutamate and tyrosine plasma levels, which were associated with gut microbiota alterations in obese patients, have been previously reported in NAFLD [40,49]. In addition, our results showed increased levels of cysteine in plasma and stool of NAFLD patients, which could lead to higher levels of hydrogen sulfide, a product of bacterial metabolism in the gut that could cause inflammation, and consequently gut health alterations. As a matter of fact, *Fusobacterium* is one of the hydrogen sulfide-producing-bacterial genus that has been reported at higher abundancy in subjects with liver diseases [50,51].

Stool samples showed increased concentrations of xanthine among NASH patients, even after BMI-adjustment. Higher levels of xanthine, which is implicated in purine metabolism, have been reported in cirrhosis and hepatocellular carcinoma [52], suggesting that this increment could be associated with DNA damage and carcinogenesis [53]. Furthermore, gut microbiota disorders have been related to alterations in nucleotide and purine metabolism [54,55]. Consequently, our results suggest that this metabolite could be an early microbiome-derived indicator of progression of disease to more severe stages of liver damage.

Correlation analysis between significant variables showed three blocks of metabolites that correlate with each other: plasma PCs and LysoPCs, plasma triglycerides and stool metabolites. Our results also showed that plasma PCs and LysoPCs negatively correlate with the rest of the significant identified metabolites. Taking together these results with those obtained from quantitative analysis, it is possible to hypothesize that there are two groups of metabolites: (1) those compounds that seem to be disease risk factors, like plasma TGs and stool metabolites, with higher concentrations in subjects with NAFLD, especially in those with NASH, and (2) those metabolites which may have a protective effect, such as plasma PCs and LysoPCs with decreased levels relative to HV.

Moreover, our results showed a positive correlation between plasma concentration of eicosenoic acid, FA(20:1) and the GG genotype of SNP rs738409 in PNPLA3 gene. This gene encodes the patatin-like phospholipase domain-containing protein 3 which has hydrolase activity toward TGs and the G allele (I148M) seems to be a loss of function mutation promoting lipids accumulation in hepatocytes [56]. In contrast to our results, it has been reported that this risk allele was not associated with metabolic changes [57]. However, this study only included patients between 34-49 years old and with fatty liver, but not necessarily diagnosed with NAFLD. Based on previous reports [34,41], a possible explanation for the encountered relationship could be speculated. Dysbiosis may contribute to FAs generation in the bowel and absorption due to increased gut permeability. Moreover, it has been described that insulin resistance, a risk factor for NAFLD, may increase adipose tissue lipolysis elevating FAs transportation to the liver, that can be detected in the bloodstream [34,41].

Our results showed that measurement of a few plasma metabolites are sufficient to discriminate HV from NAFLD patients, and more interestingly, SS from NASH patients with high accuracy (AUROC=1.00). These results are superior to those reported by Zhou et al. where a model based on clinical variables, PNPLA3 genotype, lipidomics and metabolomics data gave an AUROC=0.866 to discriminate NASH from non-NASH subjects. Moreover, our CLINICAL model, based just on PNPLA3 genotype and BMI (AUROC=0.73) is comparable to the previously described NASH Clin Score model, based on PNPLA3 genotype and other clinical data (AUROC=0.778) [39]. With the exception of the STOOL model for discriminating HV from NAFLD (AUROC=0.81), the majority of the metabolites-based models in our study had AUROC>0.90, constituting promising models compared to those reviewed by Wong et al. [58]. It is worth noting that the AUROC of the combined predictive model for disease was equal to the one obtained based only on plasma metabolites, indicating that this model could significantly simplify daily clinical screening and surveillance of NAFLD or NASH to an easy peripheral blood extraction, compared to the biopsy which is actually considered an imperfect “gold standard” method.

Although our study characterized the profile of a wide range of metabolites in HV and NAFLD patients, the low number of samples represents a limitation which does not allow us to make robust associations between the obtained results and causality of disease. Another possible limitation of our study could be the kit used for metabolomics. The MxP Quant 500 kit (Biocrates Life Sciences AG, Innsbruck, Austria) was developed for human plasma samples and it is suitable for use with fecal samples; which could possibly explain why most of the metabolites were detected in plasma rather than in stool samples, and that plasma metabolites showed the strongest associations and diagnostic accuracy. Moreover, the total of stool metabolites identified in our study was 67% higher than that reported in the kit application note [59], probably due to the applied lyophilization process [60]. Therefore, this finding highlights the importance of prudent interpretation of fecal metabolomic data as the results obtained for stool samples in our study could differ if water content is not taken into account.

## 5. CONCLUSIONS

In conclusion, this study identified several metabolites that could be considered as potential biomarkers for the diagnosis and progression of NAFLD in Argentina. BMI and plasma levels of PC aa C24:0, FA(20:1) and TG(16:1_34:1) are sufficient to differentiate between HV and NAFLD patients with high accuracy; whereas plasma levels of PC aa C24:0 and PC ae C40:1 can distinguish between SS and NASH patients with high accuracy. Further validation studies including a larger number of patients, as well as transcriptomic analysis to establish associations between these potential new biomarkers and functional microbiome networks are needed.

## Data Availability

The datasets generated and analysed during the current study are not publicly available yet. However, data may be made available from the corresponding author on reasonable request.

## AUTHOR CONTRIBUTIONS

This study was designed, directed and coordinated by J.T. and M.M.. J.T. provided conceptual and technical guidance for all the aspects of the project. A.G., S.M., L.H., P.C., A.N., M.A. and F.O. recruited NAFLD patients and collected their clinical data and food frequency consumption data. A.J.T., and S.G. recruited healthy volunteers and collected their clinical data and food frequency consumption data. M.F.M. collaborated with data collection.

F.N.M. and J.T. collected plasma and stool samples. F.N.M., F.C. and J.G. performed sample processing and metabolomics assays tutored by F.C. and J.G.

J.T. and M.F.M. performed SNP rs738409 (PNPLA3 gene) genotyping.

Data analysis was carried out by F.N.M., A.P.S., C.M.G, N.Q. and M.R.

F.N.M and J.T. drafted the manuscript.

All authors were involved in manuscript editing and approved the version submitted for publication.

## DECLARATION OF COMPETING INTEREST

All authors declare no conflict of interests.

### ACKNOWLEDGEMENTS

The authors would like to thank the patients and volunteers for their cooperation.

## Abbreviations

ALT: alanine transaminase
AST: aspartate transaminase
AUC: Area under the curve
BMI: Body mass index
FA: Fatty acid
HV: Healthy volunteers
LysoPC: Lysophosphatidylcholine
NAFLD: Non-alcoholic fatty liver disease
NASH: Non-alcoholic steatohepatitis
PC: Phosphatidylcholine
PCA: Principal component analysis
PNPLA3: Patatin-like phospholipase domain-containing protein 3
ROC: Receiver operating characteristic
SNP: Single nucleotide polymorphism
SS: Simple steatosis
TG: Triglyceride

## SUPPLEMENTAL FIGURES LEGENDS

**Supplementary Figure 1**. Metabolites identified in each group of subjects according to the sample type. Values are shown as absolute number and percentage. HV: healthy volunteers, SS: patients with simple steatosis patients, NASH: patients with non-alcoholic steatohepatitis.

**Supplementary Figure 2**. Distribution of triglycerides (TG) concentrations (μM) among samples. Median values are shown for each sample and the dotted line shows median tendency between samples. HV: healthy volunteers, SS: patients with simple steatosis patient, NASH: patients with non-alcoholic steatohepatitis.

**Supplementary Figure 3**. Receiver operating characteristic (ROC) curves for models based on clinical data (CLINICAL), plasma metabolites (PLASMA), stool metabolites (STOOL) and the combination of all the previously mentioned variables (ALL) to predict NAFLD and NASH.

